# Are ChatGPT and Copilot Reliable for Health Education on Statistical Testing?

**DOI:** 10.1101/2024.03.08.24304007

**Authors:** Alessandro Rovetta, Mohammad Ali Mansournia

**Affiliations:** Research & Disclosure, R&C Research, Bovezzo (BS), Italy; Department of Epidemiology and Biostatistics, School of Public Health, Tehran University of Medical Sciences, Tehran, Iran

## Abstract

The introduction of Artificial Intelligence (AI) has revolutionized daily life and scientific research, with applications ranging from writing scientific articles to clinical assistance. However, the effectiveness of AI models like ChatGPT 3.5 by Open AI and Bing Copilot GPT-4 by Microsoft in explaining complex concepts such as statistical testing is a cause for concern. This study investigates the ability of these AI models to explain fundamental statistical concepts, such as P-values, confidence intervals, and surprisals, crucial to properly inform conclusions in scientific research and public health. Our results highlight significant misconceptions in both AI models’ understanding and teaching of inferential statistics. These deficiencies include the mixing of incompatible statistical approaches, the nullism fallacy, the dichotomization of (statistical) significance, the incorrect interpretation of statistical measures and concepts, and an overestimation of the role of p-values and confidence intervals. Additionally, both models lack knowledge of recent alternative statistical methods like S-values and S-intervals, showing biases similar to those present in traditional statistical approaches. Given the importance of accurate statistical understanding in various sectors and the widespread integration of AI in decision-making processes, urgent intervention by OpenAI and Microsoft is necessary to update their platform databases. It is essential to align AI knowledge with the latest developments in scientific research to ensure the reliability of generated results. Collaboration with organizations such as the American Statistical Association is recommended to facilitate this process. In conclusion, this scenario underscores the need for immediate corrective action by the developing companies of such platforms. Indeed, only through continuous updates and improvements can we ensure that AI can contribute positively to scientific and technological progress.

## 1. Introduction

### Background

Artificial intelligence (AI) is rapidly transforming both our everyday lives and the landscape of scientific research. Its influence extends from seemingly mundane tasks, like improving the clarity and structure of scientific manuscripts, to the profound realm of clinical practice, where AI algorithms are being used to diagnose diseases and recommend treatment plans [1-3]. Some of the most promising health applications lie in medical imaging analysis (e.g., identifying subtle patterns and anomalies that might escape the human eye) and drug discovery and development (e.g., analyzing vast troves of biomedical data to identify promising drug targets and predicting how potential drug candidates might interact with the human body). Moreover, this infiltration of AI into the scientific sphere is not limited to professional researchers. Students and the general public are increasingly utilizing pre-trained generative language models to navigate the complexities of medical information. Whether it’s seeking clarity on COVID-19 symptoms or deciphering the intricacies of statistical tests like the Student’s t-test, AI-powered language models are empowering individuals to take a more active role in their health and education [4]. However, the integration of AI into public health and education also presents several challenges. Alongside the well-known black-box issue - making it extremely difficult to understand the methods by which an AI arrives at a certain outcome - one major concern is the potential for bias in AI algorithms [5, 6]. These biases can stem from the data and general knowledge used to train the algorithms, which may inadvertently reflect human prejudices.

### AI biases in statistical testing

In light of the above considerations, the authors of this work have questioned the current capabilities of this technology to provide adequate explanations regarding one of the most controversial and misleading topics in the scientific history of the last 100 years: statistical testing. As widely and repeatedly pointed out by leading authorities in the field, careless use of P-values and confidence intervals constantly undermines the credibility of scientific conclusions on which important decisions for public health are based [7-10]. Among these are the approval of therapeutic treatments or the implementation of anti-epidemic strategies. In particular, the main common issues are the following: i) the improper mixing of two mathematically and statistically incompatible approaches, namely (neo) Fisherian (where the P-value is sometimes improperly used as a graded measure of evidence against the null hypothesis) and Neyman-Pearson (where the P-value is used in an arbitrary and somewhat ritualistic binary decision-making fashion), ii) nullism, i.e., focusing solely on the null hypothesis of no effect, iii) the dichotomization of statistical significance (i.e., significant vs. non-significant), iv) confusion between statistical relevance and practical relevance (also known as “magnitude fallacy”), and v) lack of control over background assumptions. Therefore, this paper aims to investigate the impact of these potential cognitive and knowledge biases on the educational effectiveness in the field of statistical testing of two commonly used AI assistants: ChatGPT 3.5 from the non-profit organization OpenAI (Microsoft group) and Bing Copilot GPT-4 from Microsoft Corporation [11]. The scientific goal is to both assess the quality of the content provided by the two AI – essential from an infodemiological perspective - and to provide a collection of useful examples on the subject.

## 2. Methods

### 2.1 Data collection

In order to assess the knowledge of ChatGPT 3.5 and Bing Copilot GPT-4 on statistical testing, we devised the following series of questions: 1) What is a P-value? 2) What is a confidence interval? 3) What is a S-value? 4) What is a surprisal interval? Since the concept of surprisal interval was introduced very recently and the knowledge of ChatGPT 3.5 is updated to January 2022, the latter question was only posed to Bing Copilot GPT-4. All questions were asked sequentially to ensure continuity of topics. The responses from the two artificial intelligences were collected and copied into the main manuscript without any modifications to ensure maximum transparency. The survey was conducted between February 29 and March 7, 2024.

### 2.2 Procedure

Each response from the two AIs has been evaluated in light of the most recent and well-established evidence on statistical testing. The longer and more content-rich responses have been divided into parts (e.g., part 1, part 2, etc.); each of these has been individually commented upon. Where the AIs did not understand the request or showed a poor understanding of the question concerning the terms used, alternative, more explicit questions have been proposed. These have been fully reported in the Results section.

## 3. Results

### 3.1. Open AI ChatGPT 3.5

#### 3.1.1 What is a P-value?

##### ChatGPT 3.5 answer part 1

“*A p-value, or probability value, is a statistical measure used in hypothesis testing to determine the strength of evidence against the null hypothesis. In hypothesis testing, the null hypothesis represents a default assumption or a claim that there is no effect or no difference between groups or conditions being compared. The alternative hypothesis typically represents the researcher’s claim or the possibility of an effect or difference existing*.”

##### Comment 1

This first part of the response mistakenly mixes two mathematically incompatible approaches: the original Fisher approach (1922) and the Neyman-Pearson approach (1933). Indeed, in optimal conditions, the late-Fisherian divergence P-value (1958) can be used as an approximate, graded index of the strength of statistical evidence against a mathematical hypothesis (e.g., the null hypothesis of 0 difference between two mean values). Although Fisher himself initially defined a “convenient, arbitrary” significance threshold α = 0.05, the smaller the P-value, the greater the refutational statistical evidence. In contrast, the decision P-value by Neyman and Pearson is merely an information-free index adopted in the arbitrary criterion “P < α: reject the null hypothesis in favor of the alternative, P ≥ α: inability to reject the null hypothesis.” The only inferential information comes from numerous experiments, each conducted under the same ideal, equivalent conditions (a result impossible to achieve in practice when dealing with soft sciences like medicine) concerning the same target population (an ambiguous concept, as emphasized by Fisher, 1958) [12, 13].

##### ChatGPT 3.5 answer part 2

“*The p-value quantifies the probability of observing a test statistic as extreme as, or more extreme than, the one actually observed, under the assumption that the null hypothesis is true. In simpler terms, it tells you how likely it is to get the observed results if the null hypothesis is true*.”

##### Comment 2

This explanation is partially correct in a neo-Fisherian sense, although it omits a fundamental aspect that is the main cause of errors and uncertainty: the P-value is thus readable if and only if all background assumptions are true (e.g., data normality in parametric tests, sufficiently representative samples, functioning research tools, competence and transparency of researchers, etc.) [12-14]. Failure to meet such conditions can completely distort the interpretability of the P-value for that scope. Additionally, it should be clarified that by “observed result,” we mean the value of the test statistic and not the scientific phenomenon or its actual effect.

##### ChatGPT 3.5 answer part 3

“*A low p-value (typically below a predetermined significance level, often denoted by α, commonly set at 0.05) suggests that the observed results are unlikely to have occurred under the null hypothesis. This leads to the rejection of the null hypothesis in favor of the alternative hypothesis, indicating evidence for a significant effect or difference. Conversely, a high p-value suggests that the observed results are likely to occur under the null hypothesis, leading to the failure to reject the null hypothesis*.”

##### Comment 3

These considerations are completely incorrect and misleading. Firstly, the P-value is mathematically and epistemologically precluded from providing evidence or supporting hypotheses about what happened in the real experiment because it is calculated in a utopian world where chance - intended as the absence of causality - is the sole existing phenomenon. Since our world is made up of non-random phenomena (e.g., gravity, human wil, biochemical mechanisms), such a parallelism does not automatically hold. For the same reason, a mere P-value < α cannot indicate any practically significant effect or difference; at most, it is the researcher who must assess the relevance of the real phenomenon based on statistical information and the entire context of scientific evidence (e.g., physical, biological, clinical, psychological, etc.).

##### ChatGPT 3.5 answer part 4

“*It’s important to note that a p-value alone does not provide evidence in favor of the alternative hypothesis or the null hypothesis, nor does it quantify the size or importance of an effect. It simply helps researchers make decisions about whether to reject the null hypothesis based on the observed data. Additionally, p-values are not without controversy, and their interpretation should always be considered in the context of the study design, the specific hypotheses being tested, and other relevant factors*.”

##### Comment 4

This final recommendation, which could potentially lead to further confusion for the reader, contradicts what was previously stated by ChatGPT by correctly drawing attention to the limitations of the P-value and the importance of considering the entire scientific context. Next to this, the response again mixes up divergence P-values and decision P-values. The former can indeed help assess the incompatibility between observed data and the statistical model, but it cannot, on its own, inform any decision regarding the hypotheses under examination. The latter cannot aid in any way in evaluating rejection or acceptance: it is simply a predetermined and arbitrary decision criterion in a single experiment. Any decision regarding real phenomena must necessarily be informed by non-statistical evidence as well. This aspect is one of the cornerstones of the statistical approach since Bayes’ formulation in 1764: it is statistics to be conditional on the predetermined scientific context, not the other way around [12, 13].

#### 3.1.2 What is a Confidence Interval?

##### ChatGPT 3.5 answer part 1

“*A confidence interval is a range of values derived from sample data that is likely to contain the true population parameter with a certain level of confidence. It provides a way to quantify the uncertainty or variability associated with estimating population parameters based on sample statistics*.”

##### Comment 5

These considerations are nonsensical within both the (neo-)Fisherian and Neyman-Pearson approaches. Firstly, in the former context, the interval (1-α)*100 is improperly termed “confidence” as it expresses no statistical or any other kind of confidence (it only contains all hypotheses that, according to the adopted statistical model, have a P-value > α) [7, 15, 16]. Secondly, such an interval is mathematically precluded from estimating both the uncertainty and the variability associated with parameter estimation because i) such uncertainty also encompasses non-statistical aspects, and ii) it is not mathematically structured for this purpose. At best, its width can serve to assess statistical uncertainty (the larger the interval, the larger the statistical uncertainty) conditionally on the absence of non-random sources of uncertainty (e.g., experimental errors, bias, confounding, etc.). In contrast, in the Neyman-Pearson context, the individual confidence interval contains no information about confidence in parameter estimation merely based on - or regarding - the result obtained in the single study. Indeed, inference occurs through “coverage” over numerous (infinite) consonant executions (each under the same utopian conditions mentioned above): on average, when dealing with continuous data, (1-α)*100% of these intervals will (ideally) contain the true value (but we will never know which ones exactly) [7, 17].

##### ChatGPT 3.5 answer part 2

“*Sample Data: Researchers collect a sample from a population of interest. This sample data is used to estimate population parameters, such as the population mean or population proportion. Calculation: Using statistical methods, a confidence interval is calculated based on the sample data. The most common method involves using the sample mean (for estimating the population mean) or the sample proportion (for estimating the population proportion) and applying a formula that takes into account the sample size and the variability of the data*.”

##### Comment 6

What is written could be reasonable. However, in general, it is a serious overstatement that one can estimate the parameters of a population based on a single sample due to all the sources of uncertainty that can intervene (even unbeknownst to us). It would be more accurate to assert that properly sampling from the target population is the starting point for an inference process, which consists of numerous well-conducted independent studies. Furthermore, it is essential to emphasize - since it is a widely overlooked aspect - that inference does not only concern the generalization of the characteristics of a sample to the reference population. For example, by modeling the annual mortality from 2010 to 2019 of an entire population through linear regression, the standard errors associated with the analysis - related to the goodness of fit of the data - can be used to make inferences regarding the model’s forecast for 2020. In particular, this is one of the methods to obtain information on the comparison between observed mortality and expected mortality to assess the impact of a certain external factor (e.g., COVID-19). Specifically, this is referred to as “counterfactual analysis” since it compares what happened with what would have been expected (i.e., what could have happened).

##### ChatGPT 3.5 answer part 3

“*Interpretation: The resulting confidence interval is expressed as a range of values, often accompanied by a confidence level, which represents the probability that the interval will contain the true population parameter. For example, a 95% confidence interval for the population mean of a certain variable means that if we were to take many samples and compute a confidence interval for each sample, about 95% of those intervals would contain the true population mean*.”

##### Comment 7

The first part of the response is wrong. It is not true - even when assuming all background hypotheses to be valid (utopian scenario) - that a confidence interval of the form (1-α)*100% will contain the true value in (1-α)*100% of cases. In particular, the single already calculated interval either contains or does not contain the true value with 100% certainty! It must be clear that frequentist-inferential statistics is not designed to model our ignorance about a single event that has already occurred; instead, it is structured to predict - in the utopian scenario - hypothetical frequency patterns over entire sets of numerous experiments [7]. In simpler words, even within the utopian scenario, the probability of winning the bet “the confidence interval contains the true value” is never known unless a “sufficiently large” number “N” of statistically equivalent studies is reached (indeed, the probability of winning becomes (1-α)*100% only as “N” tends to infinity). The second statement is consistent with the Neyman-Pearson approach (especially with the late Neyman rearrangement), although it is culpably omitted that what is said works if and only if all underlying assumptions are true in each of the studies conducted (and, let’s reiterate once again, it is almost impossible for them to be perfectly true in even one of these). Finally, it should be specified that, in any case, that 95% pertains to the entirety of intervals printed in numerous studies and does not provide specific information on which of these will contain the true value and which will not [7].

##### ChatGPT 3.5 answer part 4

“*Inference: Researchers can use the confidence interval to make inferences about the population parameter. For example, if a confidence interval for the population mean does not include a certain value, we can infer that the population mean is unlikely to be that value*.”

##### Comment 8

These statements are incorrect. As already mentioned, the individual confidence interval is both mathematically and epistemologically precluded from providing inferential information about the parameter of interest (the total underlying uncertainty is too big). As easily demonstrable (supplementary file), even assuming absolutely ideal conditions (normality and absence of distortions), the value (1-α)*100% is always greater even than the probability of observing a future estimator within the associated confidence interval.

##### ChatGPT 3.5 answer part 5

“*Confidence intervals provide a more informative and nuanced understanding of the population parameter compared to point estimates (e.g*., *sample mean or proportion) because they account for sampling variability. They also allow researchers to assess the precision of their estimates and the uncertainty associated with them. Common confidence levels used include 90%, 95%, and 99%, although the choice of confidence level depends on the specific study and the desired level of certainty*.”

##### Comment 9

This is completely wrong and misleading. A confidence interval cannot provide information about, nor consider all factors that contribute to sampling variability since the sampling process also depends on non-statistical aspects that the model does not assess (e.g., the researcher’s skill and attention in designing and conducting the procedure, the correct calibration of research instruments, etc.). Similarly, a confidence interval is mathematically precluded from providing information about the precision of estimates and the associated uncertainty because the model through which it is calculated assumes the absence of non-random uncertainties as an axiom! At most, in the context of Fisher, the width of the interval can be understood as a measure of mere statistical uncertainty, assuming that the latter is the sole type of uncertainty at play (an aspect impossible to perfectly guarantee in practice) [18].

#### 3.1.3 What is a S-value?

##### ChatGPT 3.5 answer 1

“*As of my last update in January 2022, there isn’t a widely recognized statistical measure referred to as an “S-value.” It’s possible that it could be a term introduced or popularized after that time, or it could be a specific term used in a particular field or context. If “S-value” refers to a concept introduced after my last update, I would need more context or information about its definition and usage to provide a meaningful explanation. If it’s a term specific to a certain field or discipline, providing additional context would help in understanding its meaning and significance*.”

##### Comment 10

In this case, ChatGPT demonstrates a substantial ignorance in using the term “S-value” to identify the surprisal value. Therefore, we have retried with the following question. Thus, we asked: “*What is a surprisal value?*”

##### ChatGPT 3.5 answer 2

“*The term “surprisal value” is often used in the context of information theory and probabilistic models to quantify the amount of surprise or unexpectedness associated with an event or outcome. It measures how much information is gained when an event occurs. Surprisal is typically calculated as the negative logarithm of the probability of the event: Surprisal = -log2(P), where: Surprisal is the amount of surprise associated with the event. P is the probability of the event occurring. In this formula, the base of the logarithm (in this case, 2) determines the units of surprisal. When the base is 2, the unit of surprisal is a “bit” (binary digit), and surprisal quantifies the amount of information gained in bits. The intuition behind surprisal is that events with low probabilities have high surprisal values because they are more surprising when they occur, while events with high probabilities have low surprisal values because they are less surprising. Surprisal values are often used in fields such as information theory, machine learning, and natural language processing to measure uncertainty, quantify information content, and make predictions based on probabilistic models*.”

##### Comment 11

Since the response rightly encompassed a wide variety of sectors, we posed a more specific question. Thus, we asked: “*Can the surprisal value be used instead of the P-value? If yes, how?*”

##### ChatGPT 3.5 answer 3

“*The surprise value, or surprisal, serves a different purpose than the p-value in statistical inference. While both measures are related to probabilities, they have distinct interpretations and applications. Surprisal: Surprisal quantifies the amount of surprise or unexpectedness associated with an event or outcome. It is calculated as the negative logarithm of the probability of the event. Surprisal values are often used in information theory and probabilistic models to quantify the amount of information gained when an event occurs. P-value: The p-value, or probability value, is a statistical measure used in hypothesis testing to determine the strength of evidence against the null hypothesis. It quantifies the probability of observing a test statistic as extreme as, or more extreme than, the one actually observed, under the assumption that the null hypothesis is true. A low p-value suggests that the observed results are unlikely to have occurred under the null hypothesis, leading to rejection of the null hypothesis in favor of the alternative hypothesis. While both surprisal and p-values involve probabilities, they have different interpretations and applications. Surprisal measures the unexpectedness of an event, while p-values assess the evidence against a specific hypothesis. In statistical inference, the p-value is commonly used to make decisions about hypotheses, such as whether to reject or fail to reject a null hypothesis. Surprisal, on the other hand, is not typically used in this manner in traditional statistical hypothesis testing. Therefore, while surprisal and p-values both provide information about probabilities, they serve different purposes and are not directly interchangeable in statistical inference*.”

##### Comment 12

ChatGPT’s response is rather vague, confused, and at times erroneous. Besides repeating the same mistakes from previous responses regarding the P-value, it refers to the S-value as an indicator of the surprise (unexpectedness) of the actual event and deems it mathematically disconnected from the P-value. Both of these considerations are completely false since, in the context of statistical testing, the S-value is calculated as the base-2 logarithm of the P-value and is mathematically precluded from referring to the probability of the actual event for the same exact reason as the P-value (i.e., they both live in a separated world of pure chance) [16, 19]. Furthermore, it is untrue that the S-value and the P-value serve different purposes, as both can be used to inform the same conclusions and interpret the results of the same statistical test. At most, the main differences lie in the cognitive aspects related to their interpretation [20].

### 3.2 Bing Copilot GPT-4

#### 3.2.1 What is a P-value?

##### Bing Copilot GPT-4 answer

“*A p-value is a statistical measure used to validate a hypothesis against observed data. It quantifies the probability of obtaining the observed results, assuming the null hypothesis is true. Let’s break it down: Null Hypothesis (H0): In any statistical test, we start with a null hypothesis. For most tests, this hypothesis assumes that there is no relationship between the variables of interest or that there is no difference among groups. For instance, in a two-tailed t-test, the null hypothesis states that the difference between two groups is zero. Alternative Hypothesis (H0 or HA): The alternative hypothesis represents the opposite scenario. It suggests that there is a difference, relationship, or effect between the variables being studied. Calculating the p-value: When we perform a statistical test (such as a t-test or ANOVA), we obtain a test statistic (like t-value or F-value). The p-value is derived from this test statistic. It tells us how likely we are to observe the data we have if the null hypothesis were true. A small p-value indicates strong evidence against the null hypothesis, while a large p-value suggests that the observed data could have occurred by random chance. Interpreting the p-value: If the p-value is less than a chosen significance level (commonly 0.05), we reject the null hypothesis. This means we have evidence to support the alternative hypothesis. If the p-value is greater than the significance level, we fail to reject the null hypothesis. In other words, we don’t have enough evidence to conclude that there’s a significant effect. Remember, the p-value is not a guarantee of truth; it simply helps us make informed decisions about hypotheses based on the data we have*.”

##### Comment 13

These considerations are entirely similar to those made by ChatGPT 3.5, i.e., they erroneously involve the blending of Fisher and Neyman-Pearson approaches, the nullism and significance dichotomization fallacies, and the exaggeration of evidential states and scientific/epistemological possibilities of the P-value. Therefore, we refer the reader to comments 1-4. Specifically, the most serious error made here is the confusion between the mathematical hypothesis (e.g., difference in mean values equal to 0) and the scientific hypothesis (e.g., no practical difference between the placebo group and the treated group).

#### 3.2.2 What is a Confidence Interval?

##### Bing Copilot GPT-4 answer

“*A confidence interval (CI) is a statistical concept that provides a range of values within which we expect an estimate to fall if we were to repeat an experiment or resample the population. Let’s delve into the details: Definition: A confidence interval is constructed around a point estimate (such as a mean or proportion) and consists of an upper and lower bound. These bounds represent the range of values where we are confident the true parameter lies. Confidence Level: The confidence level (often expressed as a percentage) indicates how often we expect the confidence interval to contain the true parameter. For instance: A 95% confidence interval means that if we were to create many such intervals from different samples, approximately 95 out of 100 intervals would include the true parameter. The confidence level is typically denoted by 1 - α, where α (alpha) is the significance level used in statistical tests (e.g*., *α = 0.05 for a 95% confidence level). Factors Affecting Width of CI. Sample Size: Larger samples lead to narrower confidence intervals because they provide more precise estimates. Variability in the Sample: Greater variability results in wider intervals. Confidence Level: Higher confidence levels (e.g*., *99%) yield wider intervals. Example: Imagine you survey 100 Brits and 100 Americans about their weekly television-watching habits. Both groups have an average of 35 hours watched per week. However, the British group shows more variation in their viewing hours, while the Americans’ habits are more consistent. Even though both groups have the same point estimate (average hours watched), the British estimate will have a wider confidence interval due to the greater data variation. Remember, confidence intervals help us communicate the uncertainty around our estimates, allowing us to make informed decisions based on statistical evidence*.”

##### Comment 14

Although the concept of coverage is addressed, Bing Copilot GPT-4 repeats some same serious interpretative errors made by ChatGPT 3.5 (the reader is referred to comments 1-9) and even adds mathematically and statistically incorrect suggestions. Indeed, the statement that larger sample sizes correspond necessarily to narrower confidence intervals is easily disproved by the following example (we call it “example 1”). Suppose we have two samples properly drawn from two normal populations. Sample S_1_ consists of the first n_1_ = 50 natural numbers (i.e., 1, 2, 3, …, 50), while sample S_2_ consists of the first n_2_ = 100 multiples of 10 (i.e., 10, 20, 30, …, 1000). The arithmetic mean of the first sample is M_1_ = 25.5, its standard deviation is SD_1_ = 14.6, and the associated standard error of the mean is SE_1_ = SD_1_/(n_1_^0.5^) = 2.1. The 95% confidence interval is then given by (25.5 -1.96*2.1, 25.5 + 1.96*2.1) = (21.5, 29.5). Following the same procedure for the second sample, with M_2_ = 505, SD_2_ = 290, a 95% CI = (448, 562) is obtained, much wider than the first despite the double size (n_2_ = 2*n_1_). Similarly, it is false that greater variability corresponds necessarily to wider confidence intervals. In fact, the primary index of data variability is the sample variance (the square of the sample standard deviation). Considering two samples properly drawn from two normal populations with equal mean (M) and standard deviation (SD), the associated standard error of the mean SE = SD/(n^0.5^) decreases with sample size. Therefore, in this specific context and regardless of variability, the larger sample among the two will have narrower confidence intervals because (1-α)*100% = (M - zα*SE, M + zα*SE). A more general and formal proof is provided in the supplementary file. Furthermore, unless the same test in the same study is considered (which should be clearly stated), it is not true that a 99% interval is necessarily larger than a 95% interval. Taking back example 1, the 99% confidence interval of the first sample would be (20.2, 30.8), i.e., much narrower than the 95% interval of the second study, i.e., (448, 562). Finally, the conclusive recommendation regarding the use of confidence intervals to make informed decisions based on statistical evidence is much less appropriate than ChatGPT 3.5’s recommendation to integrate such “evidence” into the overall scientific context.

#### 3.2.3 Can the Surprisal Value Be Used Instead of the P-value? If Yes, How?

##### Bing Copilot GPT-4 answer

“*Certainly! Let’s explore the relationship between surprisal value and the p-value in statistical hypothesis testing. P-Value: The p-value is a fundamental concept in hypothesis testing. It quantifies the evidence against the null hypothesis. Specifically, it represents the probability of observing data as extreme as (or more extreme than) the observed data, assuming the null hypothesis is true. If the p-value is small (typically less than a chosen significance level, such as 0.05), we reject the null hypothesis in favor of the alternative hypothesis. Surprisal Value: The surprisal value, also known as surprisal, measures the unexpectedness of an event. It is based on information theory and quantifies how surprising or informative an outcome is relative to our expectations. In terms of probability, surprisal is the negative logarithm of the probability of an event: (S(x) = -\log(P(x))). Larger surprisal values correspond to more surprising events. Using Surprise Instead of P-Value: While surprisal and p-values serve different purposes, they are related. In some cases, surprisal can provide an alternative perspective to p-values. Instead of directly comparing p-values, we can compare surprisal values. If an observed outcome has a high surprisal value, it suggests that the outcome is unexpected and provides evidence against the null hypothesis. Conversely, a low surprisal value indicates that the outcome aligns with expectations. Practical Considerations: Surprisal is not commonly used as a replacement for p-values in standard statistical practice. P-values have well-established interpretations and guidelines for decision-making. Surprisal lacks the same level of standardization. However, researchers interested in information theory or seeking an alternative perspective may explore surprisal-based analyses. In summary, while surprisal and p-values are related, they serve different purposes. P-values remain the standard in hypothesis testing, but surprisal can offer an intriguing viewpoint for those interested in unexpectedness and information content*.”

##### Comment 15

Once again, we notice a detailed compendium of all the errors previously discussed regarding the P-value (see comments 1-4). Nevertheless, in this case, the misleading indications are even more perverse in the part where Copilot GPT-4 emphasizes (erroneously) the existence of a well-standardized decision criterion for the P-value that the surprisal approach does not possess. The paradox of this statement lies in the fact that the S-value was introduced in statistical testing precisely to eradicate - both cognitively and operationally - the absurdities of a decision-making ritual completely detached from any concrete scientific situation (which involves practical costs, risks, benefits, and causal - or, generally, only pseudo-random mechanisms) [16]. Additionally, the central objective of introducing S-values in public health, i.e., replacing an unintuitive measure like the P-value with the number “S” of consecutive heads when flipping a fair coin, is not even mentioned. Therefore, although the link between S-values and P-values is better clarified from a mathematical and, in some ways, statistical point of view, Copilot GPT-4 also fails to grasp the meaning and motivations behind surprisal.

#### 3.2.4 What is a Surprisal Interval?

##### Bing Copilot GPT-4 answer 1

“*Surprisal intervals, also known as S-intervals, offer an intriguing alternative to traditional statistical methods. Let’s delve into this concept: S-values (Surprisals): Statisticians propose using S-values (also known as surprisals) instead of P-values. These S-values relate the statistical relevance of an event to the number of consecutive heads when flipping an unbiased coin. In other words, S-values quantify how surprising or unexpected an outcome is, considering the context of random chance. The formula for calculating S-values is: [S\text{-value} = -\log_2(\text{p-value})] where the p-value represents the probability of observing the data under the null hypothesis. By adopting S-values, researchers can better understand the significance of their findings without relying solely on arbitrary thresholds. Surprisal Intervals (S-intervals): Building upon this concept, surprisal intervals (S-intervals) extend the idea of confidence intervals. Instead of categorizing outcomes as statistically significant or non-significant, S-intervals assess outcomes in terms of being more or less surprising than specific values. These intervals provide a range of values within which the observed data falls, considering the element of surprise. Researchers can use S-intervals to evaluate how surprising their results are relative to various hypotheses. In summary, S-values and surprisal intervals offer a fresh perspective on statistical inference, emphasizing surprise rather than rigid significance thresholds. This approach encourages more intuitive and nuanced interpretations of research outcomes. For more details, you can refer to the full paper titled “S-values and Surprisal intervals to Replace P-values and Confidence Intervals” by Alessandro Rovetta*.”

##### Comment 16

What is reported by Copilot GPT-4 is quite accurate, although there are some slightly cryptic passages and, as in previous cases, the ideal conditions in which all of this makes sense are culpably omitted. In order to remove any doubt, we examined for the interpretation of a concrete example. Thus, we asked: “*How do you interpret the following surprisal interval? 4-I = (−5, 5)*.”

##### Bing Copilot GPT-4 answer 1

“*The surprisal interval you’ve provided is denoted as [4-I = (−5, 5)]. Let’s break down its interpretation: [4-I]: This notation indicates the surprisal interval for a specific event or outcome. The “4” in the notation refers to the event of interest. Interval Limits: The lower limit of the interval is -5. The upper limit of the interval is 5. Interpretation: The surprisal interval represents a range of values within which the observed data falls, considering the element of surprise. Specifically, for the event associated with “4,” the data can vary between -5 and 5 while still being consistent with the hypothesis or model being tested. If an observed value falls within this interval, it is considered not surprising relative to the expected outcome. In summary, the surprisal interval [4-I = (−5, 5)] provides a measure of how surprising or unexpected the data associated with event “4” is, considering the context of random chance. Values within this interval are consistent with the hypothesis, while values outside it would be more surprising*.”

##### Comment 17

The explanation provided mixes elements that are literally invented (e.g., the undefined concept of “event of interest”) with potentially correct but still very vague or unclear explanations. Specifically, Copilot GPT-4 does not seem to have understood that the number “4” is the S-value (S = 4), and therefore, the associated interval (−5, 5) represents the set of hypotheses that are less surprising than 4 consecutive heads compared to the obtained statistical result (assuming the utopian scenario and that the coin is unbiased) [19]. Furthermore, it introduces erroneously and very dangerously a new form of dichotomization where a value falling within said interval should be considered non-surprising. No! Simply put, in the utopian scenario, all hypotheses contained in an S-interval are less surprising than “S” consecutive heads when flipping a fair coin as assessed by the adopted test. The last Copilot sentence is certainly the most accurate, although it contains a serious flaw: the values inside the interval are not “consistent” in an absolute sense but “more consistent” relative to certain hypotheses (in our case, to the two hypotheses whose S-value is approximately 4, namely h = 5 and h = -5). This means that, under the ideal conditions mentioned above and according to the chosen test, a hypothetical statistical effect h* = 3 would be less surprising than the hypotheses h = -5 and h = 5 compared to the observed statistical effect; or, alternatively, the hypothesis h* = 3 is less surprising than the hypotheses h = -5 and h = 5 compared to the observed statistical effect (i.e., the hypothesis h* = 3 is less surprising than 4 consecutive heads compared to the statistical effect found in the experiment).

## 4. Discussion

As of March 2024, both ChatGPT 3.5 and Bing Copilot GPT-4 have demonstrated substantial unreliability concerning the teaching of frequentist inferential statistics and related statistical testing. In particular, both artificial intelligences carry with them the cognitive distortions and interpretative errors that have afflicted much of the scientific community for over a century [6]. Among the main mistakes, we highlight the mixing of mathematically and epistemologically incompatible approaches of Fisher and Neyman-Pearson, the tendency to reconsider the null hypothesis of no effect (nullism fallacy), the dichotomization of statistical significance, the omission of interpretability conditions of P-values (background assumptions), gross overestimation of the roles and possibilities of concepts such as statistical significance and confidence intervals, attribution of improper statistical properties to confidence intervals, as well as the presentation of conflicting information. Alongside this, the aforementioned AIs lack structured knowledge even on very recent alternative methods such as S-values and S-intervals, instead applying the latter the same biases related to P-values and confidence intervals. Such a scenario is consistent with some reports in previous literature on human biases transmitted to artificial intelligence systems [6]. Therefore, based on the well-documented evidence on statistical testing, immediate intervention by Open AI and Microsoft is required to update their products’ databases, aligning basic knowledge with the state of the art of scientific knowledge in this regard [7-22].

Indeed, the need for urgent corrective action is based on a series of critical considerations. First of all, the importance of the correct understanding and application of statistical testing cannot be underestimated, as these concepts are essential within scientific and decision-making analysis in a wide range of fields, from economics to medicine, from engineering to psychology [22]. Misinterpreting statistical results can lead to incorrect conclusions and harmful decisions, with significant consequences at both individual and societal levels. Secondly, the advent and widespread adoption of artificial intelligence-based technologies, such as ChatGPT and Bing Copilot, increase the responsibility of the companies developing them to ensure the accuracy and reliability of the results generated by such platforms. In an era where AI is increasingly integrated into daily life and choices, it is crucial not only for intelligent technologies to be informed by updated and as accurate knowledge as possible but also for there to be a rapid method to implement any important corrections. Finally, keeping AI databases aligned with the latest developments in the field of statistical testing (or other relevant sectors) is necessary to ensure that these technologies continue to be at the forefront and relevant in the rapidly evolving environment of scientific and technological research.

## Conclusions

The inadequacy shown by ChatGPT 3.5 and Bing Copilot GPT-4 in understanding and teaching inferential statistics and statistical testing is a cause for concern and requires immediate intervention from the companies developing them. Only through a joint commitment to updating and improving the knowledge of these platforms can we ensure that AI continues to play a positive and reliable role in scientific and technological progress. Collaborations with organizations such as the American Statistical Association are recommended in this regard.

## Data Availability

All data produced in the present work are contained in the manuscript

