## Supplementary File for "Are ChatGPT and Copilot Reliable for Health Education on Statistical Testing?"

### Appendix 1

Alessandro Rovetta, Mohammad Ali Mansournia

March 2024

#### 1 Supplementary file

##### 1.1 Confidence intervals: probability of containing future estimates (normal distributions in ideal conditions)

$$x \sim N(\mu, \sigma),\ y \sim N(\mu, \sigma) \mid \mathrm{E}(x) = \mathrm{E}(y) = \mu$$

$$Var(\delta) = s_\delta^2 = Var(x - y) = Var(x) + Var(y) = s_x^2 + s_y^2$$

$$p(\delta, s_\delta) = \frac{1}{s_\delta\sqrt{2\pi}} \exp\Big[ -\frac{(\delta - h)^2}{2s_\delta^2} \Big] = \frac{1}{\sqrt{2\pi(s_x^2 + s_y^2)}} \exp\Big[ -\frac{(x - y - h)^2}{2(s_x^2 + s_y^2)} \Big]$$

$$s_y = ks_x = ks \Longrightarrow p_k(\delta, s) = \frac{1}{s\sqrt{2\pi(1 + k^2)}} \exp\Big[ -\frac{(\delta - h)^2}{2(1 + k^2)s^2} \Big]$$

$$P_k(h - zs < \delta < h + zs) = \frac{1}{s\sqrt{2\pi(1 + k^2)}} \int_{h-zs}^{h+zs} \exp\Big[ -\frac{(\delta - h)^2}{2(1 + k^2)s^2} \Big] d\delta$$

$$\lim_{k \to 0} p_k(\delta, s) = p(\delta, s_\delta)$$

| $z$ | k | $P$ |
|---|---|---|
| 1.96 | 0 | 0.95 |
| 1.96 | 0.5 | 0.92 |
| 1.96 | 1 | 0.83 |
| 1.96 | 2 | 0.62 |
| 1.96 | 3 | 0.46 |

#### 1.2 Confidence intervals: relationship between width, sample size, and variability (normal distributions in ideal conditions)

$$x = x_i \sim N(\mu_x, \sigma_x),\ y = y_i \sim N(\mu_y, \sigma_y) \mid \mathrm{E}(x) = \mu_x,\ \mathrm{E}(y) = \mu_y$$

$$\bar{\omega} = \frac{1}{n_\omega}\sum_{i=1}^{n_\omega}\omega_i,\ s_\omega = \sqrt{\frac{1}{n_\omega - 1}\sum_{i=1}^{n_\omega}(\bar{\omega} - \omega_i)^2},\ \bar{s}_\omega = \frac{s_\omega}{n_\omega}$$

$$(1-\alpha)\cdot 100\% = (\bar{\omega} - z_\alpha \bar{s}_\omega,\ \bar{\omega} + z_\alpha \bar{s}_\omega) \mid \Delta_\omega = \bar{\omega} + z_\alpha \bar{s}_\omega - \bar{\omega} - (-z_\alpha \bar{s}_\omega) = 2 z_\alpha \bar{s}_\omega$$

$$\Delta_x = 2z_\alpha \frac{s_x}{\sqrt{n_x}}\ \tilde{\mathcal{R}}\ \Delta_y = 2z_\alpha \frac{s_y}{\sqrt{n_y}} \iff \frac{s_x}{\sqrt{n_x}} - \frac{s_y}{\sqrt{n_y}} = \frac{s_x}{\sqrt{n_x}} - \frac{k s_x}{\sqrt{l n_x}} =$$
$$\frac{s_x}{\sqrt{n_x}}\left(1 - \frac{k}{\sqrt{l}}\right) \mathcal{R}\ 0 \iff 1\ \mathcal{R}\ \frac{k}{\sqrt{l}}$$

$$k > \sqrt{l} \implies \mathcal{R} : <,\ k < \sqrt{l} \implies \mathcal{R} : >$$
